# Characteristics and phenotypes of a COPD cohort in Uganda

**DOI:** 10.1101/2023.05.11.23289559

**Authors:** Patricia Alupo, Levicatus Mugenyi, Winceslaus Katagira, Kayongo Alex, Nalunjogi Joanitah, Trishul Siddharthan, John R Hurst, Bruce Kirenga, Rupert Jones

**Affiliations:** Makerere University Lung Institute, Kampala, Uganda; Department of Medicine, Makerere University College of Health Sciences; University of Miami; UCL Respiratory, University College London, London, UK; Plymouth Marjon University, Plymouth, UK; Medical Research Council, Uganda Virus Research Institute and London School of Hygiene and Tropical Medicine, Entebbe Unit, Uganda

## Abstract

**Introduction:** Chronic Obstructive Pulmonary Disease (COPD) is a heterogeneous condition with varied clinical and pathophysiologic characteristics. Although there is increasing evidence that COPD in low- and middle-income countries may have different clinical characteristics from that in high income countries, little is known about COPD phenotypes in these settings. We describe the clinical characteristics and risk factor profile of a COPD population in Uganda.

**Methods:** We cross sectionally analyzed the baseline clinical characteristics of 323 COPD patients aged 30 years and above who were attending two national referral outpatient facilities in Kampala, Uganda between July 2019 and March 2021. Logistic regression was used to determine factors associated with spirometric disease severity.

**Results:** The median age was 62 years; 51.1% females; 93.5% scored CAT >10; 63.8% mMRC >2; 71.8% had wheezing; 16.7% HIV positive; 20.4% had a history of pulmonary tuberculosis; 50% with blood eosinophilic count >3%, 51.7% had 3 or more exacerbations in the past year. Greater severity by GOLD stage was inversely related to age, (aOR=0.95, 95%CI=0.92, 0.97), and obesity compared to underweight (aOR=0.25, 95%CI=0.07, 0.82). Regarding clinical factors, more severe airflow obstruction was associated with SPO2<93% (aOR=3.79, 95%CI=2.05, 7.00), mMMRC≥2 (aOR=2.21, 95%CI=1.08, 4.53), and a history of severe exacerbations (aOR=2.64, 95%CI=1.32, 5.26).

**Conclusion:** COPD patients in this population had specific characteristics and risk factor profiles including HIV and TB meriting tailored preventative approaches. Further studies are needed to better understand the pathophysiological mechanisms at play and the therapeutic implications of these findings.

**Key messages:**

- **What is already known on this topic.** COPD is a heterogeneous condition with the greatest burden in LMICs yet there is limited understanding of disease characteristics in this setting.
- **What this study adds** A cohort of patients with COPD recruited in hospital clinics in Uganda showed a high burden of disease with frequent exacerbations – 86% were GOLD category D. The cohort had a high exposure to biomass smoke and only 38% were past or present smokers.
- **How this study might affect research, practice, or policy** There is need for more research into effective strategies to prevent and treat COPD in LMICs - it cannot be assumed that guidelines derived in high-income countries will apply.

## Background

Chronic Obstructive Pulmonary Disease (COPD) is a heterogeneous lung condition characterized by chronic respiratory symptoms (dyspnea, cough, expectoration and/or exacerbations) due to abnormalities of the airways (bronchitis, bronchiolitis) and/or alveoli (emphysema) that cause persistent, often progressive, airflow obstruction (1). COPD is the third leading cause of death and Disability Adjusted Life Years (DALY’s) globally, with 90% of these deaths occurring in Low- and Middle-Income Countries (LMICs) (2)(3) (4) (5). In 2019, 3.3 million deaths and 74.4 million DALYs were attributed to COPD (6). Furthermore, the COPD burden is projected to keep increasing due to increased aging of the population and continued exposure to risk factors (7). COPD is a complex syndrome with numerous pulmonary and extra-pulmonary components. It is not yet clearly known how different patterns of exposures affect the clinical presentation, physiology, imaging, response to therapy, lung function decline and survival (8)(9). Recent research in COPD has been aimed at better understanding the heterogeneity across different COPD patient groups and phenotyping COPD patients for better understanding of this condition (10).These phenotypes are also aimed at categorizing those with similar clinical characteristics and their response to treatments, to better guide management approaches (11)(12)

The most widely known risk factor for the development of COPD is tobacco smoke exposure, and most of the knowledge about COPD has been based on tobacco exposed populations. In sub–Saharan Africa, the prevalence of COPD is high, with a pooled prevalence of 8%, and associated risk factors include increasing age, smoking, history of pulmonary tuberculosis and biomass smoke exposure(13)(14). Significantly, indoor, and outdoor air pollution, occupational exposures and infections have also emerged as risk factors (15)(16). These varied risk factors have a different pathway of effect on the lung compared to tobacco smoke(17). Unique host factors including nutritional status, high HIV prevalence and previous tuberculosis (TB) also contribute to the risk of developing the airflow obstruction that is characteristic of COPD in LMICs. Little is known about how the different patterns of noxious exposures and other risk factors affect the pathological and clinical phenotypes. Additionally, there is scarcity of data concerning COPD patient populations from LMICs. Van Gamert et al, following a cross sectional study in Uganda, found that people with COPD were younger (30-40 years) compared to those in High Income Countries (HIC’s) who are usually older than 40 years, and presented more commonly with wheeze compared to COPD in high income countries (18). Soriano and colleagues found that 70% of COPD in HICs was attributed to tobacco smoking, while environmental exposures accounted for 60% of COPD in LMICs (19).These findings allude to potential variation between COPD in HICs and LMICs and there is insufficient data from LMICs to provide robust management advice(20). Further region-specific research is needed to bridge this knowledge gap and better guide management approaches. This is especially true for LMICs where resources are limited and the burden of COPD is high. We have described the clinical characteristics at baseline of a population with COPD in Uganda, with substantial biomass exposure.

## Methods

### Study design and setting

We enrolled a cohort of 323 COPD patients, who presented at the chest clinic outpatient department of Mulago National referral hospital, Kiruddu referral hospital and the Lung Institute clinic, all situated in Kampala, Uganda.

The Lung Institute Clinic is a specialist outpatient chest clinic situated within the college of health of Health sciences, Makerere University, Kampala, Uganda. Given these are referral facilities, they attract patients from across the country.

We enrolled participants who were aged 30 years and above, who consented to participate in the study, and with the aim of being followed up for one year to evaluate the frequency and predictors of exacerbations among these patients. We describe the baseline characteristics of these patients. The sample size was obtained using the primary objective of the study, using the formula given by Lwanga and Lemeshow (21)

### Study procedures

We diagnosed COPD using quality assured spirometry according to ATS standards in patients who had a supporting history of dyspnea, chronic cough, sputum production and a history of exposure to risk factors for the disease(22).

We included participants with a diagnosis of COPD aged 30 years and above who provided informed consent. We excluded participants who had active pulmonary TB (PTB) and other co-morbid diseases likely to affect participation or outcomes, such as lung cancer and asthma as deemed by the investigational team. Notably, the COVID-19 pandemic and restrictions slowed patient enrollment and follow up due to the travel restrictions and fear of hospital settings.

Spirometry was conducted by trained technicians, using the Vitalograph Pneumotrac spirometer with Spirotrac software (vitalograph Ltd;Buckingham, United Kingdom). It measured the forced vital capacity (FVC), forced expiratory volume in one second (FEV1), the FEV1/FVC ratio and flow volume curves. We obtained three acceptable manouvers in accordance with ATS/ERS guidelines (23). A COPD diagnosis was defined as a post-bronchodilator FEV1/FVC ratio below 0.7 together with appropriate symptoms, such as dyspnea, chronic cough, sputum production, wheezing/chest tightness, recurrent chest infections in the context of a history of risk factors, following GOLD guidelines. GOLD staging was performed using post-bronchodilator spirometry data, an assessment of dyspnoea using the modified MRC dyspnoea scale and the exacerbation history. Exacerbations were defined as the number of times patients experienced worsening of respiratory symptoms that warranted additional medication, and this often resulted in a visit to a health facility or pharmacy.

At enrolment, all consenting participants underwent a respiratory focused clinical evaluation using a case report form (CRF) to collect data on demographics, symptoms, exposures to outdoor and indoor pollutants, maternal exposures, childhood medical history, exposure to known COPD risk factors, tobacco smoking, biomass smoke exposure, psychosocial issues including dysfunctional breathing, co-morbidities (allergies, gastro esophageal reflux disease (GERD), diabetes mellitus (DM)), impact of COPD on the person’s life using the COPD Assesment Test (CAT) and Clinical COPD Questionaire (CCQ), inhaler technique, vital signs and respiratory system physical signs. Baseline venous blood and induced sputum samples were collected.

### Ethical considerations

Ethical approval for was obtained from the Mulago Hospital Research and Ethics Committee MHREC 1451, and the Uganda National Council for Science and Technology (HS 2483).

### Statistical analysis

Patients’ baseline characteristics were summarized using proportions for categorical data; and mean (standard deviation) or median (interquartile range) for continuous variables. The study outcome was the patients’ GOLD stage defined as binary outcome derived by grouping gold stages I and II into one category (mild-moderate) and gold stages III and IV into another category (severe-very severe). Biomass exposure was cross tabulated with tobacco smoking history and results presented in a Venn diagram. A Chi-square test was used to determine the association between biomass exposure and tobacco smoking. Logistic regression was used to determine factors associated with disease severity. All factors with p values less than 0.2 at a bivariable analysis (simple logistic regression) were subjected to a multivariable logistic regression, checked for multicollinearity and model building done using Akaike Information Criteria (AIC). Results from the final fit are presented as adjusted odds ratios (OR) with 95% confidence intervals (CI). For all associations, a p value of less than 0.05 was considered statistically significant. STATA version 15 was used for all analyses.

## Results

We screened 457 participants with spirometry who had presented to the health facilities with cough, wheeze, dyspnea, chest tightness and/or sputum/phlegm production. 323 were eligible and consented to participate in the study, while 134 failed screening. Reasons for failing screening include, a lack of bronchodilator reversibility on spirometry (35%), normal spirometry findings (45%), having restrictive pattern on spirometry (8.2%), failure to complete the spirometry procedure (9.7%), uncontrolled blood pressure (0.7%), and 1.5% of them simply declined to participate in study (Fig1). We enrolled a total of 323 participants, with a spirometry diagnosis of COPD between July 2019 and March 2021(Table 1). The mean age of the participants was 62.9 years, and 51.1% were females. The symptom burden of the participants at presentation was high, with 93.5% having a COPD Assessment Test (CAT) score of 10 and higher, and 63.8% scoring 2 and above on the Modified medical research council (mMRC) dyspnea score (Table 1).

**Figure 1.**
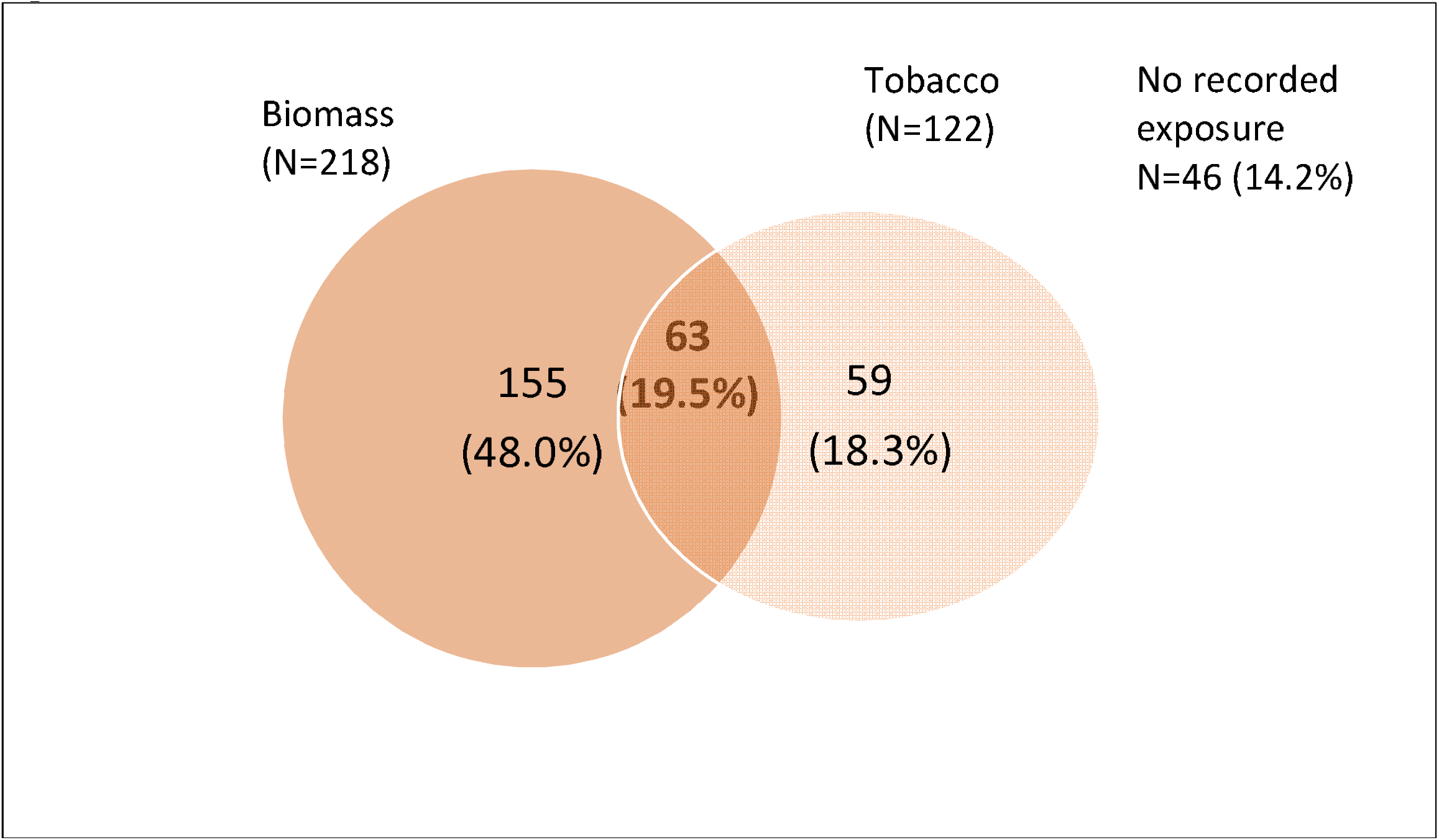
Venn diagram showing numbers and percentages of 323 participants with smoking and biomass exposure.

**Table 1.**
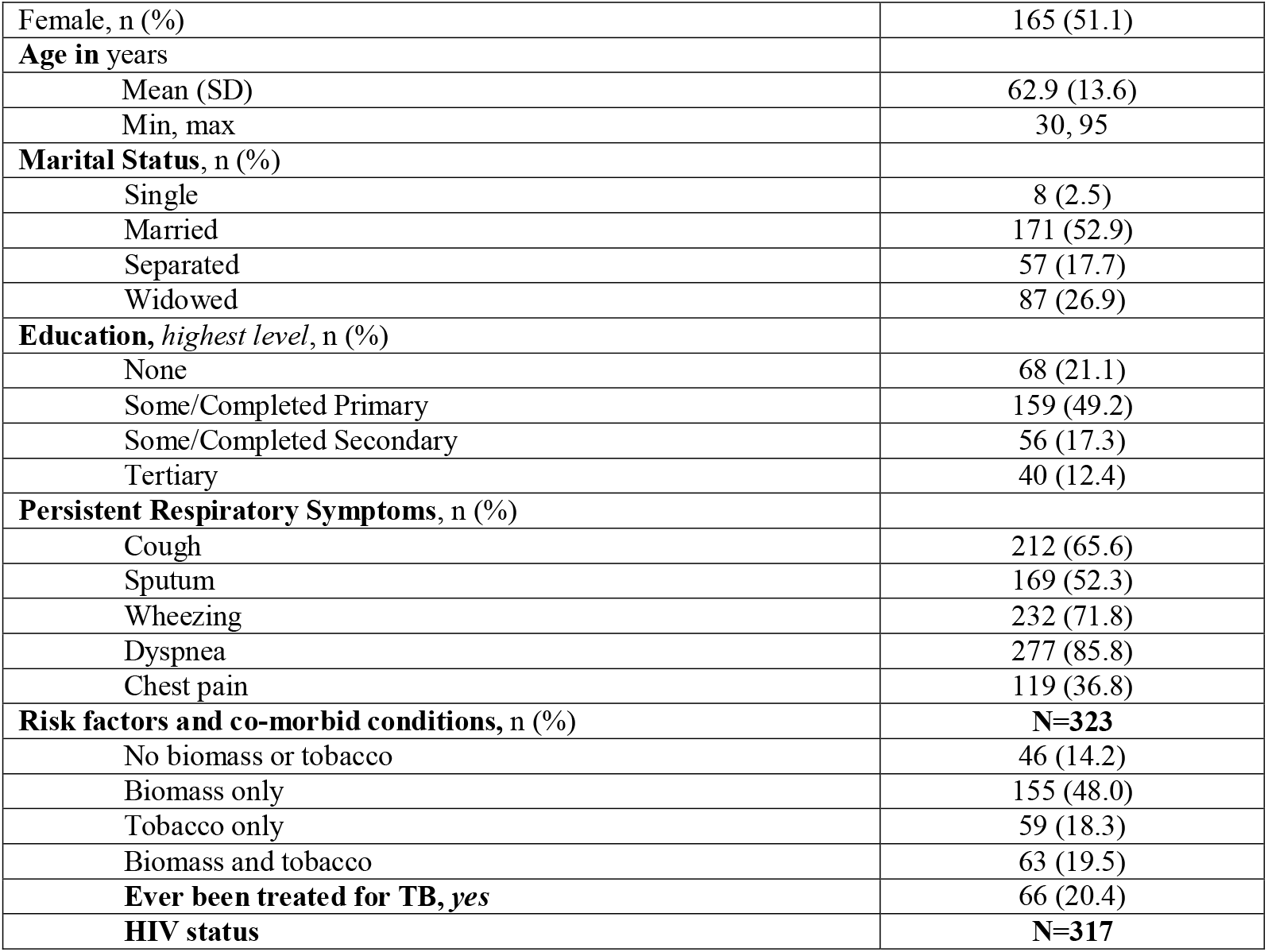
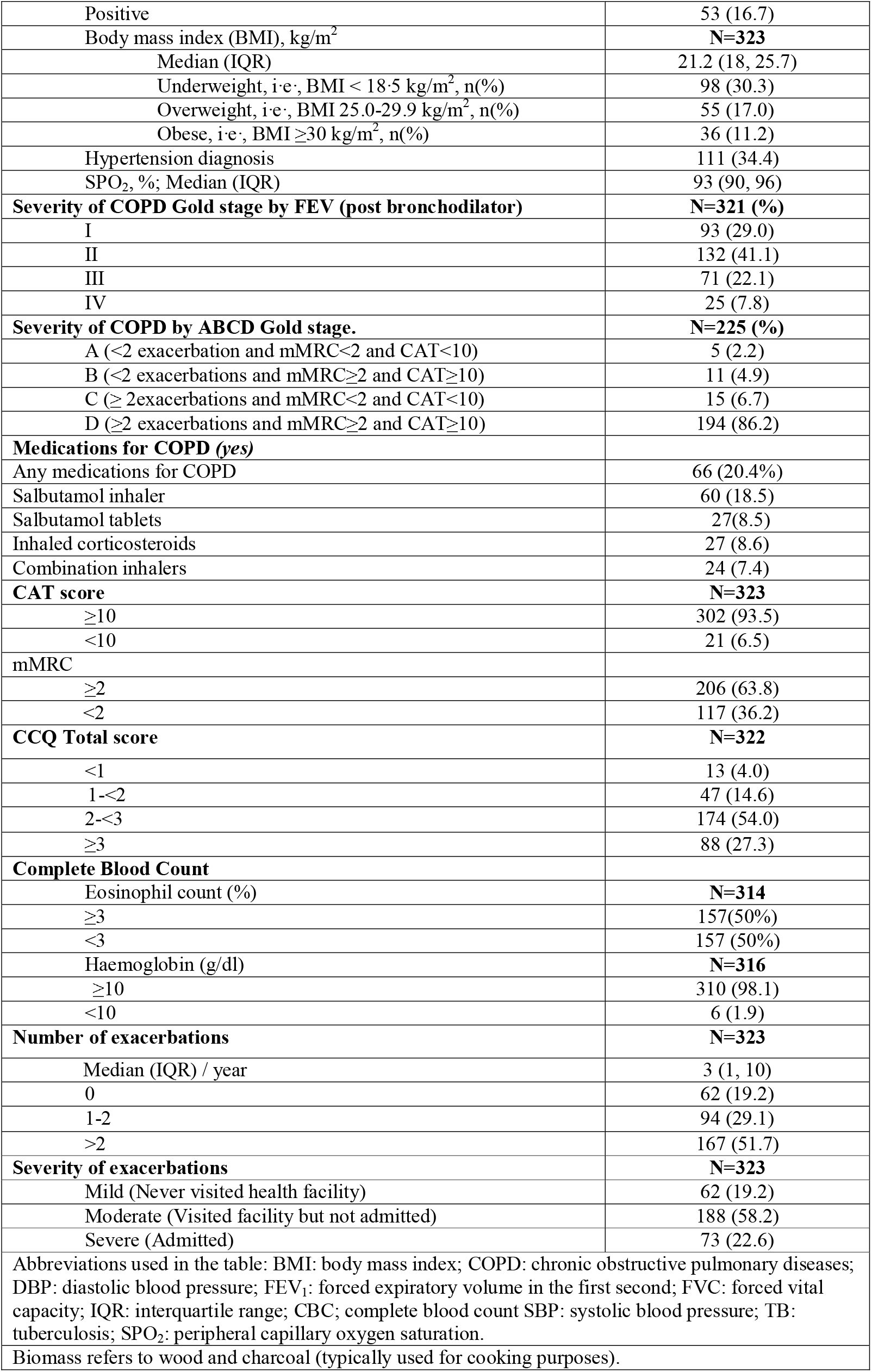
Baseline characteristics of COPD patients presenting to a pulmonary center in Kampala (N=323)

Participants in GOLD stage III and IV were more often hypoxic, underweight, and had a higher symptom burden. (Table 2) They also reported a history of more frequent and more severe exacerbations compared to those in GOLD I/II. Interestingly, participants in GOLD stage III and IV were younger than those in GOLD stage I and II. On assessment of co-morbidities, hypertension was the most prevalent co-morbidity with 34% of the participants having a history of hypertension. 22% of the participants reported a history of PTB, and there was a high prevalence of HIV in this population, with 16.7% of the participants who were eligible and enrolled being HIV positive.

**Table 2.**
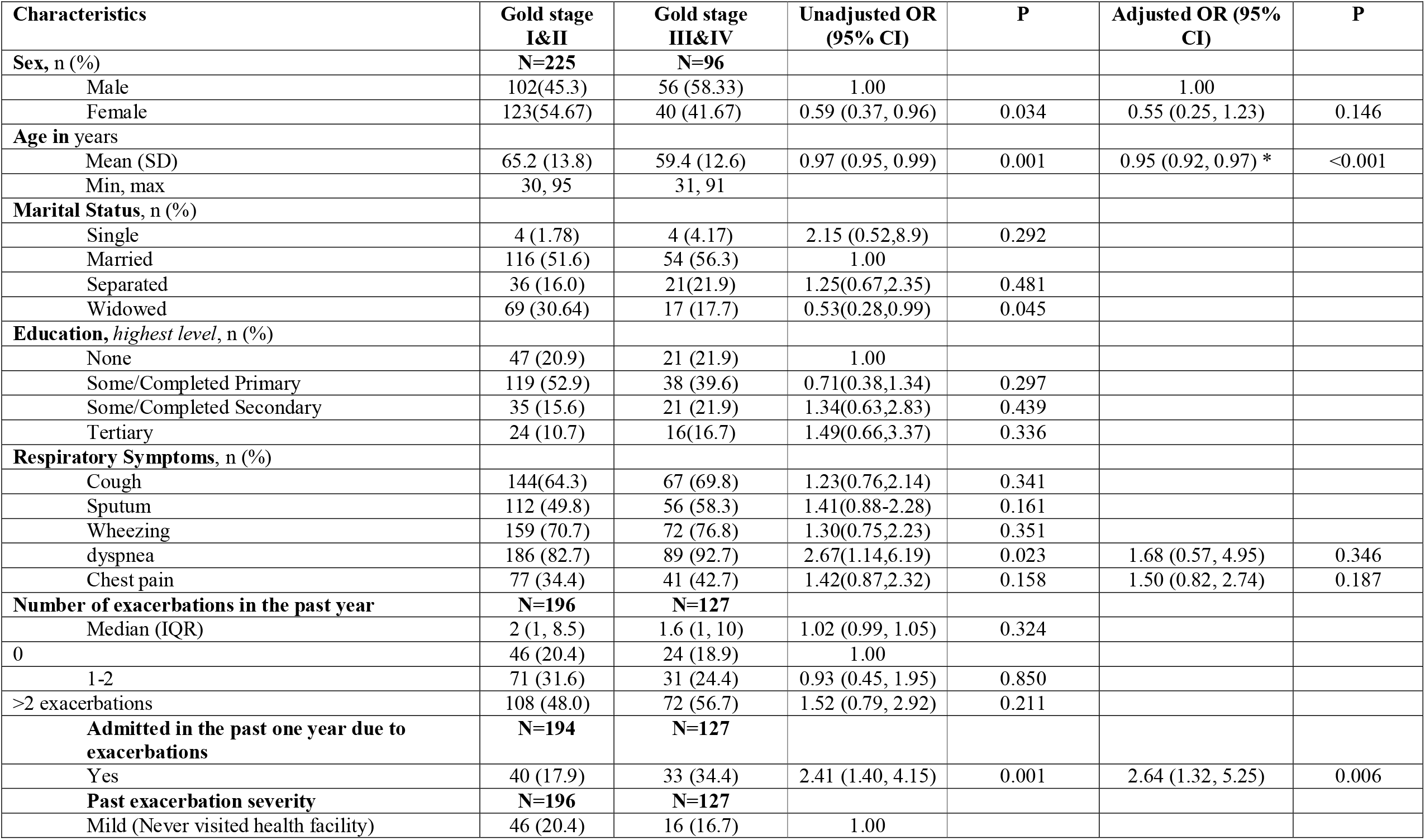
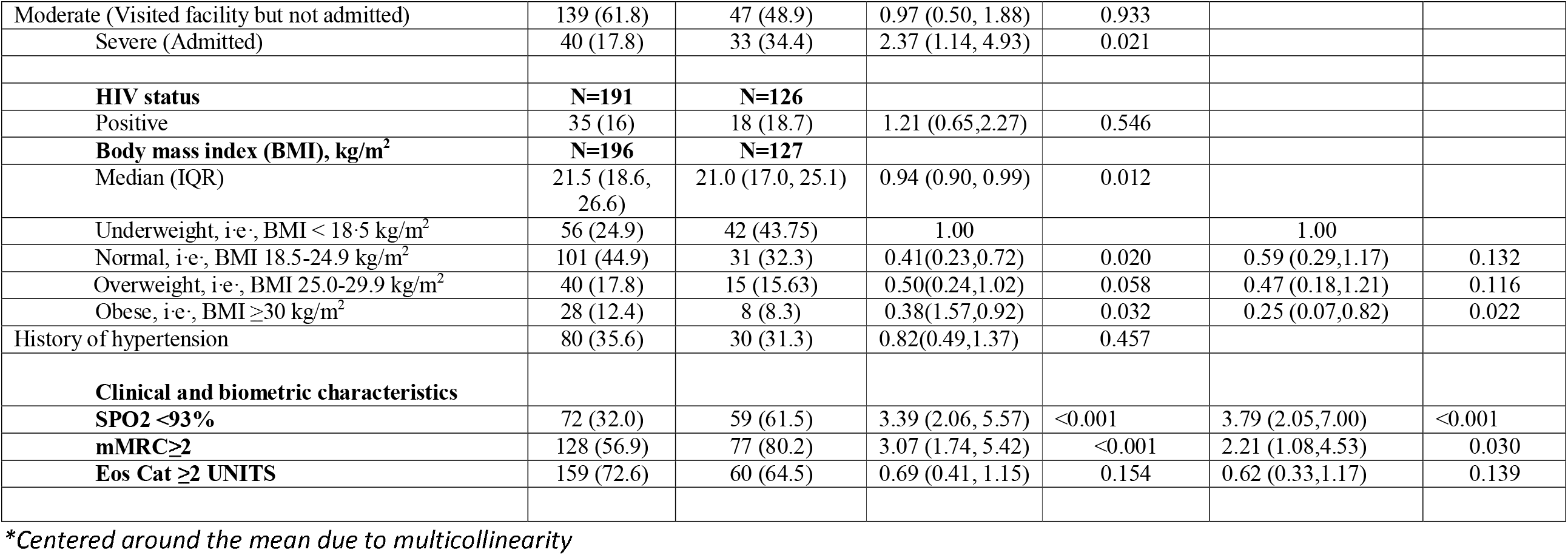
Baseline characteristics associated with disease severity defined according to GOLD stage (I & II vs III & IV) for patients presenting to a pulmonary center in Kampala.

Table 2 shows relationships between baseline participants’ characteristics and disease severity (stage III or IV versus stage II or III). The factors significantly associated with severe disease and their adjusted odds ratios, aOR (95% CI) were: age aOR=0.95 (0.92, 0.97)); exposure to biomass and tobacco smoke, that is, biomass only aOR=0.31 (0.12, 0.83), tobacco only aOR=0.32 (0.11, 0.91), and both biomass and tobacco aOR=0.29 (0.10, 0.83) (reference to no biomass or tobacco); being obese (compared to normal weight) aOR=0.25 (0.07, 0.82); SPO2 less than 93% aOR=3.79 (2.05, 7.00); number of exacerbation in the past one year aOR=1.01 (1.003, 1.01); and hospital admission in the past one year aOR=2.64 (1.32, 5.25). The factors which were associated with disease severity but became statistically insignificant after adjusting for other factors were: sex (female vs male) unadjusted odds ratio, uOR (95% CI)= 0.59 (0.37, 0.96); and severity of past exacerbations, that is, having a history of admission uOR (95% CI)= 0.59 (0.37, 0.96) compared to mild (never visited health facility).

## Discussion

In this observational study of 323 COPD patients who presented to two outpatient health facilities in Uganda, we found that the population with COPD was young, predominantly female, with patients having a high exacerbation frequency, symptom burden and a high HIV prevalence. Biomass exposure was the most ubiquitous risk factor, although tobacco smoke and pulmonary tuberculosis were also common. Dyspnea was the most prevalent symptom, followed by wheezing, but by disease severity, patients with severe disease had dyspnea as the most frequent presenting symptom, while those with mild and moderate disease presented more with cough. There was equal distribution concerning the peripheral blood eosinophil count with 50% of the participants having eosinophilia of >3%.

The older, and the obese patients had less severe airflow obstruction compared to the younger, and underweight patients. Factors associated with more severe disease were oxygen saturation <93% by oximetry, younger age, mMRC≥2, and having a history of hospitalization due to exacerbations.

The clinical characteristics of this cohort seem to be different to those seen in studies from high-income countries. It is difficult though to compare this cohort directly to cohorts from other settings. Firstly, there are differences in the health care systems, treatment availability and referral processes. Secondly there are relatively few studies published on COPD patients recruited from outpatient clinics However overall COPD populations from general population studies, those recruited from primary care report low rates of exacerbations with a majority with GOLD A. In this study we found a median annual exacerbation rate of 3. In comparison to the United Kingdom (UK) primary care, a study of over 9000 patients showed much lower rates-mean exacerbation rate was 0.89 / year. GOLD Group A: 36%, B: 19%, C: 20%, D: 25%(24). In secondary care populations Donaldson’s study of outpatients in East London the median exacerbation rate was 2.5 / year, although this relied on diary card recording which usually gives higher results (25). In the ECLIPSE study the mean rate of exacerbations was 1.2/ year [(26). In particular, the finding of 86% with GOLD category D is much higher than other populations derived from hospital patients. In a study of the combined populations in the ECLIPSE, the COPD gene and the COCOMICS cohorts: the severity distribution was group A: 32%, group B: 21%, group C: 10% and Group D: 37% (27). The implication of frequent exacerbations in this cohort is that past exacerbations are the best guide to future exacerbations (26). Frequent exacerbations are associated with a range of adverse outcomes and this population may be at high risk. Given the young age and female predominance, and that COPD is common disease in LMICs, urgent steps are needed to understand the prognosis and prevention / treatment strategies in global settings. After all, most of the burden and 90% of global COPD deaths occur in LMICs. Additionally, not all COPD is the same and further work is needed to better understand the pathological and clinical characteristics of the disease in LMICs. Equally, not all exacerbations are the same-some are infectious and others inflammatory, studies are needed on the etiology and prevention of exacerbations in various settings in LMICs – the pattern may be different in rural versus urban environments.

Paradoxically, in this population, we found that younger age was associated with more severe disease likely, and this could be as a result of behavioral related comorbidities in the young, that contribute to more disease severity(28). Alternatively, this may reflect a survivor effect in older people with milder disease.

The higher female predominance in this population is akin to findings by *Van Gemert et al* who had similar findings in a rural setting in Uganda, and similar to the recent global trends(29), which was not the case previously (18) (3). The higher prevalence of COPD in women in this population could be explained by the fact that in this setting, women are culturally the principal cooks in the home and as a result, likely have more prolonged and intense exposure to biomass smoke (18).

Biomass smoke was the most prevalent risk factor in this population, and this supports findings from prior studies in Uganda and as has been demonstrated by the patterns and trends of COPD in LMICs in relation to HICs (30). 14.2% of the participants reported having been neither exposed to tobacco smoke or biomass smoke. We did not exhaustively evaluate risk factors such as occupational exposures or outdoor air pollution, and these could have been the primary risk factor in the 14.2% of participants who had had neither biomass or cigarette smoke exposure in their history(31). Non cigarette risk factors are progressively significantly contributing to the global burden of COPD (32)(33). Wheezing was the second most prevalent presenting symptom after dyspnea, which suggesting and airway predominant phenotype in this population(34).

The HIV prevalence of 16.7% in this population was 3 times the current national HIV prevalence in Uganda. This is much higher than the prevalence in previous studies in Uganda, which were 10.2% and 6.2% by *Alupo et al* and *Kayongo et al* respectively, thus demonstrating a trend of an increasing prevalence of HIV in COPD patients in Uganda(35)(36). The high prevalence we observed may also be as a result of more frequent HIV diagnosis now. The high HIV prevalence we observed is not unique to this population though, as some studies have demonstrated the progressively increasing prevalence of COPD among HIV patients globally (37). This increase in COPD prevalence among HIV patients has mainly been attributed to the increasingly aging HIV patient population, resulting from better HIV control conferred by the Highly Active Anti-Retroviral Therapies (HAART), and thus more people with HIV having the opportunity to develop COPD compared to the pre-HAART era (38). It could also be explained by the pathophysiological mechanisms of HIV in the respiratory tract, predisposing the HIV patient population to chronic respiratory tract disease (39).

A history of PTB was present in 22% of participants, and thus alludes to PTB being the underlying risk factor for COPD in this patient group. This would thus place them under the COPD-1 etiotype as per the GOLD 2023 taxonomic classification. (1) This is supported by findings from studies conducted prior in this setting, evaluating the burden of COPD in both rural and urban Uganda conducted by *Kayongo et al*, and *Ddungu et al* respectively (36)(40).Similar findings have been demonstrated globally (41), and highlights the need for TB control as a strategy for alleviating the burden of COPD in this setting where the TB prevalence is high (42).

The main limitation of the study is the fact that the data was the cross-sectional design of the analysis. More robust prospective studies are needed to further support these findings.

## Conclusion

COPD patients in this Ugandan secondary-care population have specific characteristics including younger patient age, presentation with dyspnea and wheeze, frequent exacerbations and biomass smoke being the most prevalent exposure. HIV and history of pulmonary tuberculosis are common among these patients, and thus more attention should be accorded to COPD by HIV and TB healthcare providers in this setting. More research is needed to better understand the pathophysiological mechanisms at play in view of the unique risk factor profile, and how these translate to the preventative and therapeutic approaches in this COPD phenotype.

## Data Availability

All data produced in the present study are available upon reseaonable request to the authors

